# EVALUATION OF THE RELATIONSHIP BETWEEN ADVANCED MATERNAL AGE AND PREGNANCY OUTCOME: A SCOPING REVIEW

**DOI:** 10.1101/2024.03.05.24303764

**Authors:** Innocent Okafor Eze, Oliver Chukwujiekwu Ezechi, Ahmadu Shehu Mohammed, Uchenna Ifeanyi Nwagha

## Abstract

**Background:** Childbirth at advanced maternal age (AMA) is increasing among women, and some studies have shown adverse outcomes. There is a need to map evidence on the subject to harness findings and identify research gaps for further studies. This scoping review aims to examine pertinent studies on AMA and its impact on pregnancy outcomes.

**Methods:** The Preferred Reporting Item for Systematic Review and Meta-Analysis (PRISMA) chart is employed for systematic data extraction. This review draws from Google Scholar, the Cochrane Library, Medline (via PubMed), and Embase (via OVID). For inclusion in this scoping review, articles must thoroughly examine and elucidate the effects, impacts, and relationships between advanced maternal age and pregnancy outcomes. A crucial prerequisite is that the articles undergo a peer-review process to ensure the reliability and credibility of the presented information. For this review, advanced maternal age is defined explicitly as women aged ≥35. However, studies focusing on women aged ≥40 are also considered, mainly if they are high quality. In terms of research methodology, both primary and secondary research will be eligible, encompassing systematic reviews and meta-analyses. This broad inclusion aims to capture a comprehensive overview of the existing literature on the subject. Furthermore, articles must be presented in the English language to facilitate a standardized and accessible analysis. This criterion ensures that language barriers do not impede the review’s ability to synthesize relevant information effectively.

**Results:** There are significant associations between advanced maternal age and poor pregnancy outcomes, even when adjusted for confounders.

**Conclusions:** The adverse pregnancy outcome due to maternal age alone may be due to placental dysfunction resulting from a relative deficiency in maternal cardiovascular adaptations to pregnancy, and this provides a window for further studies.

## Introduction

Worldwide, childbirth at an older age, also known as advanced maternal age (AMA) (defined by the International Federation of Gynecology and Obstetrics (FIGO)) as age ≥35□years at the time of expected delivery) is significantly increasing among women (1-3). The reasons are multifactorial: civilization, effective contraceptives, longer life expectancy, gender equity and equality, and more education and career prospects (4, 5).

In most civilized societies, cultural evolution has liberated women from early marriage, childbearing, and home-keeping. Furthermore, lack of suitable partners, male selection preference, negative marital experience of others, and desire for large family sizes (developing countries) are some socio-cultural factors contributing to delayed motherhood (Olabusoye et al. 2020).

Additionally, in recent years, advances in assisted reproductive therapy (ART) have challenged the traditional age-related limitations of reproduction, enabling even postmenopausal women to conceive and give birth (6). Data from the World Health Organization Multicountry Survey (WHOMCS), mainly on developing countries, reveal a prevalence of 12.3% (16% for Nigeria) of pregnancies among women aged 35 years and above between 2010 and 2011(7). This demographic shift toward delayed childbearing is becoming a primary clinical and public health concern because of the well-documented association with adverse maternal and neonatal pregnancy outcomes.

There is a need to do a scoping review of studies that addressed the subject to map evidence and identify research gaps for further studies. This scoping review mapped out the local and global prevalence of AMA and also assessed the impact of AMA on pregnancy outcomes. In addition, the review identified possible research gaps in the subject for further studies.

## Method

This study constitutes a scoping review encompassing selected studies conducted in the past decade. The review adhered to a five-step methodology following the methodological framework for scoping studies outlined by Arksey and O’Malley (2005). The process was visually represented using a PRISMA flow chart (Fig. 1) and involved the following stages: formulating the research question, identifying pertinent studies and subsequently selecting them, charting the gathered data, and ultimately summarizing and reporting the results.

**Figure 1.**
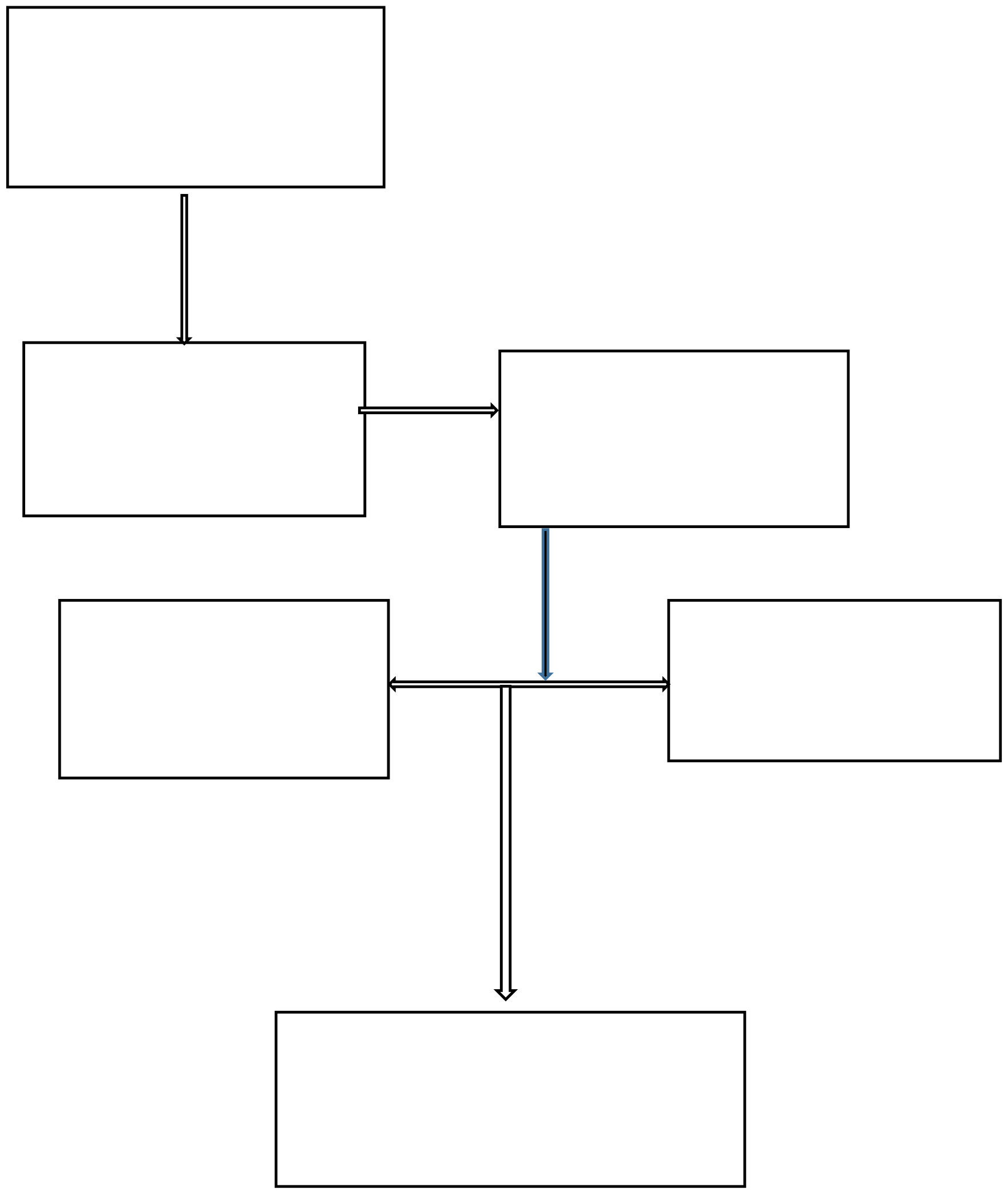
Preferred Reporting Item for Systemic Review and Meta-Analysis (PRISMA) chart.

### Review questions

1. Is there significant evidence supporting the association between advanced maternal age (AMA) and poor pregnancy outcomes?
2. In what ways does advance maternal age impact pregnancy outcomes?
3. What research gaps exist in the current literature, necessitating further studies to enhance our understanding of the relationship between advanced maternal age and pregnancy outcomes?

### Eligibility

#### Inclusion criteria

1. The article must explicitly examine the effects, impacts, and relationships between advanced maternal age and pregnancy outcomes.
2. Inclusion requires that the article undergoes a peer-review process to ensure rigor and reliability.
3. Advanced maternal age is defined as women aged ≥35, with consideration of ≥40 for high-quality studies.
4. Both primary and secondary research are eligible, encompassing systematic reviews and meta-analyses.
5. Articles must be presented in the English language to facilitate uniform understanding and analysis.
6. Studies were selected based on the quality of evidence in decreasing order of priority: meta-analysis, systematic review, prospective cohort studies with a relatively large sample size, retrospective studies with a relatively large sample size, and geographical spread.

#### Exclusion criteria

1. Editorials and perspectives, summaries of workshops, and conference abstracts.
2. Studies that do not directly address the research question.
3. Non-peer-reviewed articles.
4. Duplicate studies presenting similar information within the same environment.
5. Studies deemed of relatively poor quality.

### Information sources and search

A search was conducted across four databases: Google Scholar, Cochrane Library, Medline (via PubMed), and Embase (via OVID) to identify and retrieve relevant studies from peer-reviewed articles investigating the impact, relationship, and effects of advanced maternal age on pregnancy outcomes. The search was delimited to studies conducted within the past ten years, from 2013 to 2023.

The keywords used in the search include maternal age, advanced maternal age, older maternal age, and pregnancy outcome. Search range was for studies published between the years 2013 to 2023 (16 April 2023, last date for search). The selection process mirrors the contemporary demographic landscape pertinent to the topic at hand. However, a study outside the search range was included based on the need for inclusiveness/spread and quality.

#### Synthesis of results

In keeping with a scoping review methodology, the inclusion and exclusion criteria were defined a priori and were refined as necessary throughout the iterative screening process at the beginning, middle, and end of the screening process to ensure consistency. Articles were selected during title and abstract screening if they met the inclusion criteria

## Results

### Data charting process

This was done using the Preferred Reporting Item for Systemic review and Meta - Analysis (PRISMA) chart as shown below. (See Figure 1)

To gather evidence from local (Nigerian) studies, eight articles published on the subject in Nigeria during the specified period were assessed. From these, three pertinent studies were chosen based on their geographical coverage and quality of evidence, ensuring representation of the three main ethnic groups and regions in Nigeria: Hausa, Igbo, and Yoruba, which predominantly correspond to the Northern, Eastern, and Western parts of the country, respectively. Notably, for the Western region, a study falling outside the initial search range was included for its quality.

In the case of foreign studies, the preliminary search yielded 7,392 articles, narrowed down to 1,190 within the specified timeframe. Following the removal of non-peer-reviewed articles, grey literature, duplicates, and studies not addressing the research question, 82 articles remained. These peer-reviewed articles underwent systematic screening, with Selection based on relevance, quality of evidence, and geographical spread. The criteria for selecting studies, arranged in decreasing order of importance, included meta-analysis, systematic review, prospective cohort studies with a relatively large sample size, retrospective studies with a relatively large sample size, and geographical spread, all other factors being equal. Applying these criteria, seven studies were chosen for review, offering a global perspective with representation from Europe, North America, Asia, and the Middle East. This Selection, in addition to the three local studies, resulted in a total of ten studies included in the review.

### Charting the data

The item list used for charting data is presented in Table 1. Information on first author, year of publication, type of research, methodological design, study setting (i.e., country in which the study was performed or specifics of healthcare organization in which the study was carried out), participant characteristics such as specific patient group or professionals (where relevant) and major findings were charted.

**Table 1:**
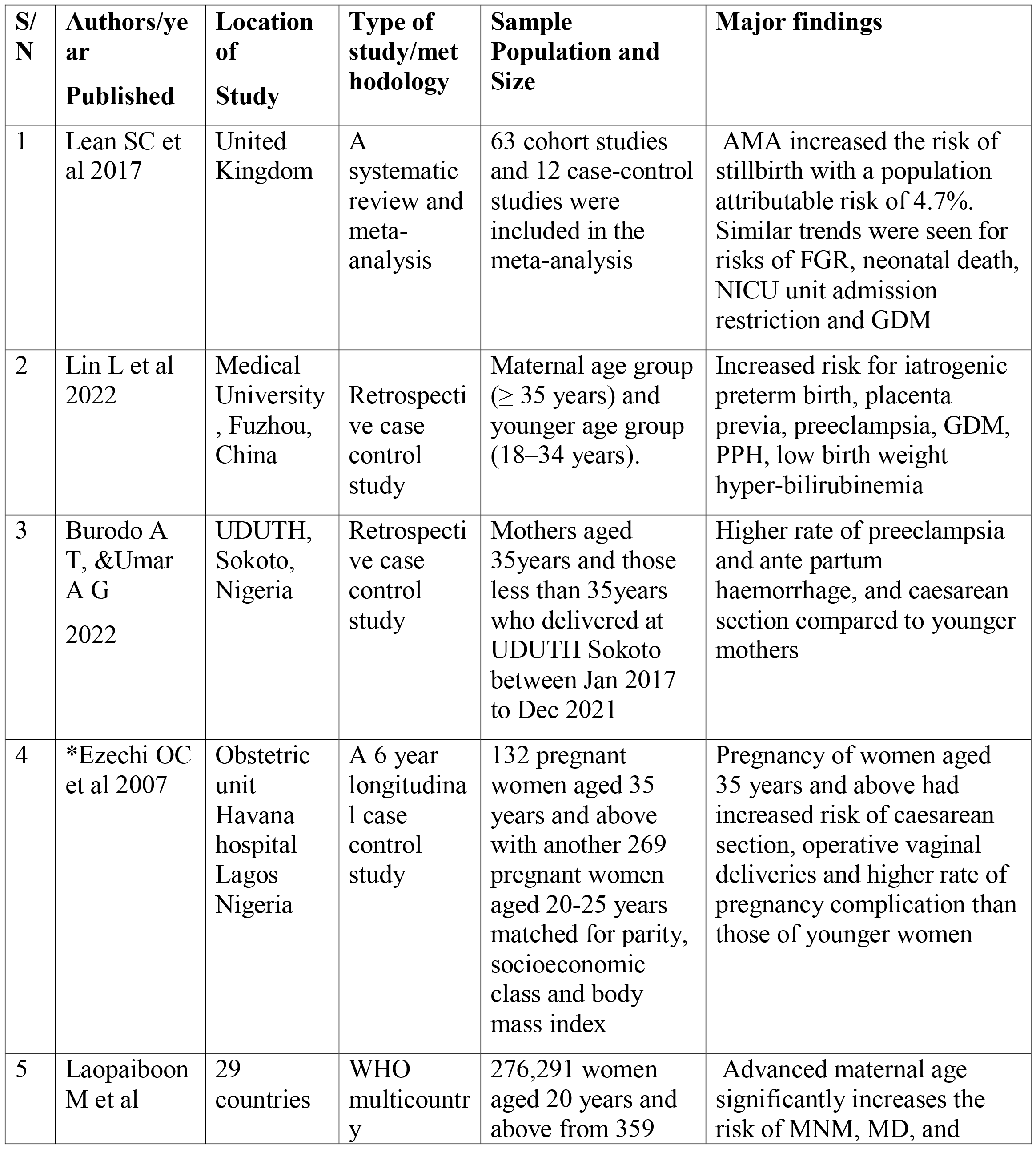
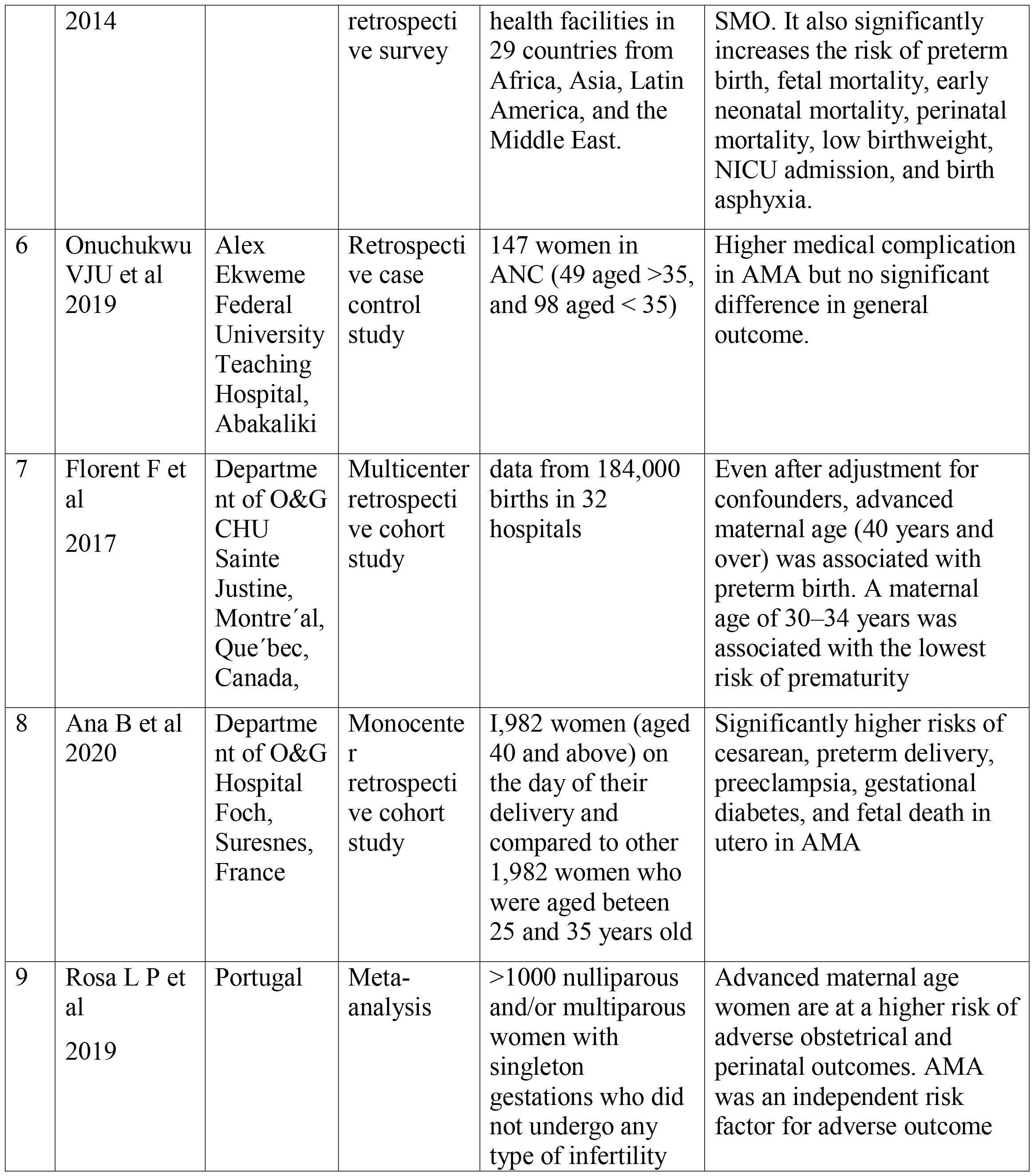
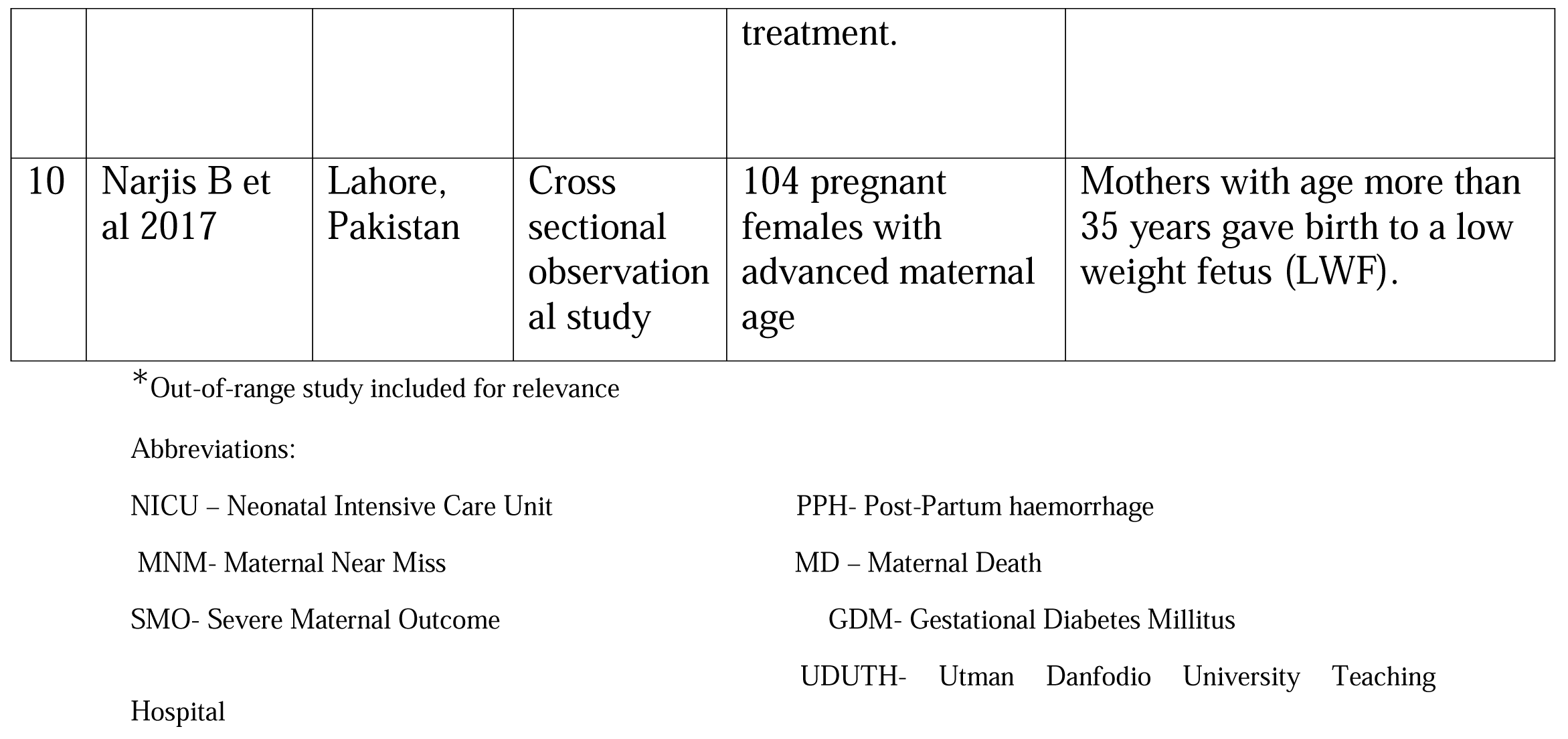
Basic characteristics of studies reviewed.

| **S/**<br>**N** | **Authors/year**<br>**Published** | **Location**<br>**of**<br>**Study** | **Type of**<br>**study/met**<br>**hodology** | **Sample**<br>**Population and**<br>**Size** | **Major findings** |
| --- | --- | --- | --- | --- | --- |
| 1 | Lean SC et al 2017 | United Kingdom | A systematic review and meta-analysis | 63 cohort studies and 12 case-control studies were included in the meta-analysis | AMA increased the risk of stillbirth with a population attributable risk of 4.7%. Similar trends were seen for risks of FGR, neonatal death, NICU unit admission restriction and GDM |
| 2 | Lin L et al 2022 | Medical University , Fuzhou, China | Retrospective case control study | Maternal age group ($\geq 35$ years) and younger age group (18–34 years). | Increased risk for iatrogenic preterm birth, placenta previa, preeclampsia, GDM, PPH, low birth weight hyper-bilirubinemia |
| 3 | Burodo A T, &Umar A G 2022 | UDUTH, Sokoto, Nigeria | Retrospective case control study | Mothers aged 35years and those less than 35years who delivered at UDUTH Sokoto between Jan 2017 to Dec 2021 | Higher rate of preeclampsia and ante partum haemorrhage, and caesarean section compared to younger mothers |
| 4 | *Ezechi OC et al 2007 | Obstetric unit Havana hospital Lagos Nigeria | A 6 year longitudinal case control study | 132 pregnant women aged 35 years and above with another 269 pregnant women aged 20-25 years matched for parity, socioeconomic class and body mass index | Pregnancy of women aged 35 years and above had increased risk of caesarean section, operative vaginal deliveries and higher rate of pregnancy complication than those of younger women |
| 5 | Laopaiboon M et al | 29 countries | WHO multicountry | 276,291 women aged 20 years and above from 359 | Advanced maternal age significantly increases the risk of MNM, MD, and |
|  | 2014 |  | retrospective survey | health facilities in 29 countries from Africa, Asia, Latin America, and the Middle East. | SMO. It also significantly increases the risk of preterm birth, fetal mortality, early neonatal mortality, perinatal mortality, low birthweight, NICU admission, and birth asphyxia. |
| 6 | Onuchukwu VJU et al 2019 | Alex Ekweme Federal University Teaching Hospital, Abakaliki | Retrospective case control study | 147 women in ANC (49 aged >35, and 98 aged < 35) | Higher medical complication in AMA but no significant difference in general outcome. |
| 7 | Florent F et al 2017 | Department of O&G CHU Sainte Justine, Montréal, Québec, Canada, | Multicenter retrospective cohort study | data from 184,000 births in 32 hospitals | Even after adjustment for confounders, advanced maternal age (40 years and over) was associated with preterm birth. A maternal age of 30–34 years was associated with the lowest risk of prematurity |
| 8 | Ana B et al 2020 | Department of O&G Hospital Foch, Suresnes, France | Monocenter retrospective cohort study | 1,982 women (aged 40 and above) on the day of their delivery and compared to other 1,982 women who were aged between 25 and 35 years old | Significantly higher risks of cesarean, preterm delivery, preeclampsia, gestational diabetes, and fetal death in utero in AMA |
| 9 | Rosa L P et al 2019 | Portugal | Meta-analysis | >1000 nulliparous and/or multiparous women with singleton gestations who did not undergo any type of infertility treatment. | Advanced maternal age women are at a higher risk of adverse obstetrical and perinatal outcomes. AMA was an independent risk factor for adverse outcome |
| 10 | Narjis B et al 2017 | Lahore, Pakistan | Cross sectional observational study | 104 pregnant females with advanced maternal age | Mothers with age more than 35 years gave birth to a low weight fetus (LWF). |

\*Out-of-range study included for relevance
Abbreviations:
NICU – Neonatal Intensive Care Unit
PPH- Post-Partum haemorrhage
MNM- Maternal Near Miss
MD – Maternal Death
SMO- Severe Maternal Outcome
GDM- Gestational Diabetes Mellitus
Hospital
UDUTH- Utman Danfodio University Teaching

### Characteristics and Selection of sources of evidence

Out of the ten studies, 3 were done in Nigeria and deliberately each selected from the North (Sokoto), East (Ebonyi) and the Southern (Lagos) part of the country. Given her multi-ethnic and cultural diversity, this is to take fair geo-cultural representation of Nigeria. The third study conducted in Lagos, Southern Nigeria was off the intended range but was selected due to study location and the prospective nature of the design. The 6 other studies reviewed were done in Europe (3), and one each from Asia, Middle East and Canada. A WHO multicounty study involving 359 health facilities in 29 countries from Africa (including Nigeria), Asia, Latin America, and the Middle East was also reviewed. Of the 3 studies from Europe (UK, Portugal and France), 2 were meta-analysis while one was a retrospective cohort study.

### Synthesis of results

Two of the studies (done in the North and Eastern part of Nigeria) were retrospective cohort in nature and focused on a comparative 5-year review of pregnancy outcomes between older (>35 age) and younger (<35 age) primigravida.

The prevalence of older primigravida was 0.2% and 0.42% in the North and the East, respectively. The Northern study noted that Igbo Christians had a significantly higher proportion of elderly primigravida compared to the Northern Muslims. Both studies conclusively found a higher rate of medical complications in pregnancy in older mothers compared to younger ones. However, whereas the study conducted in the North (8) found a significantly higher rate of antepartum hemorrhage and cesarean section in older mothers but fetal outcome was similar in both arms, the study in the Eastern Nigeria (9) did not find any significant difference in general outcome in both cohorts. The third study was conducted in Lagos, Southern Nigeria. The study conclusively found that pregnancy of women aged 35 years and above women had an increased risk of cesarean section, operative vaginal deliveries, and a higher rate of pregnancy complication than those of younger women (10).

The six other studies reviewed were done in Europe (3), and one each from Asia, the Middle East, and Canada. A WHO multicounty study involving 359 health facilities in 29 countries from Africa (including Nigeria), Asia, Latin America, and the Middle East was also reviewed. Of the three European studies (UK, Portugal, and France), 2 were meta-analyses, while one was a retrospective cohort study. The UK study (11) was a meta-analysis of 74 studies on AMA with primary outcomes as stillbirth and fetal growth restrictions and other secondary outcomes, including SGA infants, LBW infants, preterm birth, neonatal deaths, NICU admission, preeclampsia and placental abruption and GDM. Despite the large degree of heterogeneity, the extensive meta-analysis was consistent in the theme that increasing maternal age is associated with stillbirth rates FGR. Most AMA-related conditions identified in the analysis have strong biological associations with placental dysfunction, providing a logical avenue for future study. Further understanding the mechanisms underpinning this increased risk of stillbirth in AMA may provide novel tools to identify women at the highest risk of stillbirth so that intervention may be applied to improve outcomes for this high-risk population.

The other meta-analysis (4) looked at ten studies with a population of > 1000 nulliparous and/or multiparous women with singleton gestations who did not undergo any infertility treatment. Advanced maternal-age women were more likely to be overweight and have gestational diabetes and gestational hypertension. They were also more likely to undergo induced labor and elective cesarean deliveries. Furthermore, they had worse perinatal outcomes such as preterm delivery, low birth weight babies, higher rates of Neonatal Intensive Care Unit admission, and worse Apgar scores. Advanced maternal-age women had higher rates of perinatal mortality and stillbirth.

The France study (12) was a mono-center retrospective cohort study of 1,982 women (aged 40 and above) at the day of their delivery and compared to equal number of women aged between 25 and 35. Significantly higher risks of cesarean, preterm delivery, preeclampsia, gestational diabetes, and fetal death in utero were found in AMA.

In a retrospective case-control study in China in cohorts of maternal age group (≥ 35 years) and younger age group (18–34 years), findings were increased risk for iatrogenic preterm birth, placenta previa, preeclampsia, GDM, PPH, and low birth weight hyper-bilirubinemia (13).

In Pakistan (14) conducted a cross-sectional observational study of 104 pregnant females with advanced maternal age and found a significantly higher risk of low birth weight fetuses (LWF).

The Canadian study (15) was a multicenter retrospective cohort study of 184,000 births in 32 hospitals. The study found that even after adjustment for confounders, advanced maternal age (40 years and above in this study) was associated with preterm birth. A maternal age of 30–34 years was associated with the lowest risk of prematurity.

In the WHO multi-country retrospective survey of 276,291 women aged 20 years and above from 359 health facilities in 29 countries, it was found that advanced maternal age significantly increases the risk of maternal near miss (MNM), maternal death (MD) and severe maternal outcome (SMO). It also significantly increases the risk of preterm birth, fetal mortality, early neonatal mortality, perinatal mortality, low birth weight, NICU admission, and birth asphyxia.

## Discussion

### Summary of evidence

Numerous studies have investigated the effects of advanced maternal age on pregnancy outcomes. Most of these studies are retrospective, with a significant proportion conducted in high-income countries. This prevalence can be attributed, in part, to the demographic trend of delayed childbearing, which seems more pronounced in high-income nations than in countries with fewer resources. This demographic shift could contribute to the abundance of studies on advanced maternal age and pregnancy outcomes in affluent European nations, among other factors.

For instance, the 2011 WHO multi-country survey revealed significant variations in the prevalence of Advanced Maternal Age (AMA) among participating nations, ranging from 2.8% in Nepal to 31.1% in Japan (Laopaiboon et al., 2014). Interestingly, this trend is not exclusive to high-income countries, as evidenced by the same survey. Nigeria, with a 16% prevalence, surpassed countries like China (9%), Mexico (11%), and Brazil (12%) in the proportion of pregnancies in advanced age. This underscores the multifaceted factors contributing to this global phenomenon.

In the context of Nigeria, limited studies focusing on the effects of AMA on pregnancy have been conducted in the last six years, primarily adopting a retrospective approach. Moreover, these studies often narrow their focus to nulliparous women experiencing AMA. Examining the prevalence of AMA across ethnic groups in Nigeria reveals variations, with rates ranging from 0.2% in the North (Burodo & Umar) to 1.79% in the West (Ezechi et al., 2007). However, these figures may be somewhat underestimated compared to the WHO general survey.

The discrepancies arise from methodological variations. Two studies (North and East) exclusively considered elderly primigravida, omitting multiparous pregnant women who, due to a high desire for multiparity, might outnumber nulliparous women in the group. The Western study included multiparous women but used age 40 instead of 35 as the cut-off for AMA, excluding a significant proportion of the study group. Additionally, the Northern study noted that Igbo Christians had a significantly higher proportion of elderly primigravida compared to Northern Muslims. This disparity was attributed to ethnic socio-cultural differences, where Northern Muslim women, with lower formal education, tended to marry earlier than Igbo Christians who migrated to the North mainly for academic pursuits, consequently delaying childbirth to an older age.

These findings align with broader research indicating that women’s academic and career pursuits are major factors contributing to delayed childbearing. Recognizing these patterns is crucial for health policymakers, guiding planning and intervention strategies to address the implications of delayed childbearing among women. Most studies examining the effects of AMA on pregnancy outcomes consistently demonstrate a negative impact compared to younger women, holding other factors constant. Across all studies, a prevalent finding is the heightened occurrence of medical complications during pregnancy, particularly hypertensive disorders and diabetes mellitus. This observation aligns with expectations, considering the well-established correlation between advanced age and an increased risk of these medical complications during pregnancy.

Interestingly, a subset of studies (Bouzaglou et al., 2020; Lean et al., 2017; Pinheiro et al., 2019) conducted adjustments for variables such as hypertensive disorders and diabetes mellitus yet still identified a significant negative impact of AMA on pregnancy outcomes. Notably, all studies tried to exclude multiple gestation and ART pregnancies from their study populations, enhancing the precision of their findings.

Furthermore, a consistent finding across almost all studies was the significantly increased likelihood of cesarean delivery associated with AMA, particularly in elderly primigravida (Burodo & Umar; Ezechi et al., 2007; Uchenna et al., 2019). This pattern may be linked to the perception of pregnancies in advanced age as exceptionally valuable and high-risk, potentially leading to a preference for cesarean delivery over vaginal delivery. Additionally, the medical conditions more commonly associated with advanced age, which can negatively impact pregnancy, may contribute to an increased need for cesarean delivery among women with AMA. These observations underscore the multifaceted nature of the challenges posed by advanced maternal age in the context of pregnancy outcomes. The studies reviewed presented inconsistent findings regarding adverse perinatal and neonatal outcomes associated with Advanced Maternal Age (AMA). Meta-analyses and studies with diverse populations and substantial sample sizes (Laopaiboon et al., 2014; Lean et al., 2017; Pinheiro et al., 2019) consistently observed a significant association between AMA and adverse perinatal and neonatal outcomes compared to younger mothers. Conversely, local studies with relatively small sample sizes (Burodo & Umar; Ezechi et al., 2007; Uchenna et al., 2019) did not find any significant differences in fetal outcomes between the two age groups.

Lean et al. (2017) highlighted that AMA-related conditions identified in their analysis had strong biological associations with placental dysfunction, indicating a potential avenue for future research. Moreover, understanding the mechanisms underlying the increased risk of stillbirth in AMA could offer novel tools for identifying women at the highest risk, allowing interventions to improve outcomes for this high-risk population.

This review’s strengths include high-quality studies with large sample sizes incorporating systematic reviews and meta-analyses. This broad Selection of sources helps mitigate bias and enhances the reliability of the findings.

However, certain limitations should be acknowledged. A scoping review inherently lacks depth in assessing the quality of evidence. Additionally, the review is constrained by the decision to focus on the past ten years and select only a few studies for review, potentially omitting some relevant evidence.

## Conclusion and Recommendation

The comprehensive analysis of the reviewed studies consistently reveals a notable association between advanced maternal age and adverse feto-maternal outcomes. While this association is evident, it is essential to consider whether maternal age alone can entirely account for such outcomes. Placental dysfunction emerges as a plausible mediator, potentially attributed to a relative deficiency in maternal cardiovascular adaptations to pregnancy.

To further elucidate this hypothesis, it is imperative to conduct prospective studies that specifically investigate the role of placental dysfunction in the context of advanced maternal age. Such studies should investigate the intricate mechanisms underlying the observed associations, allowing for a more nuanced understanding of the interplay between maternal age, placental function, and feto-maternal outcomes. This research avenue contributes to advancing our knowledge and provides valuable insights for developing targeted interventions and healthcare strategies to mitigate the potential risks associated with advanced maternal age. Moreover, collaborative efforts between researchers, health policymakers, and healthcare professionals are crucial in facilitating the design and implementation of these prospective studies, ensuring their robustness and relevance to clinical practice.

## Data Availability

All data produced in the present study are available upon reasonable request to the authors

## Acknowledgement

The authors acknowledge the administrative support of the Commander Nigerian Navy Reference Hospital Ojo, Surgeon Capt. IB Chukwuka.

